# Exploring the interdependence between telemedicine and frugal innovation in the context of adoption and diffusion in low-resource settings: a systematic review protocol

**DOI:** 10.1101/2025.07.01.25330611

**Authors:** Christopher Adlung, Pouria Paridar, Caroline Figueroa, Christa Niehot, Nikita Kanumoory Mandyam, Cees van Beers, Saba Hinrichs-Krapels

**Affiliations:** Faculty of Technology, Policy, and Management, Delft University of Technology, The Netherlands; Medical Library, Erasmus Medical Center Rotterdam, The Netherlands; Equitech Collective; Faculty of Technology, Policy, and Management, Delft University of Technology, The Netherlands & Erasmus Medical Center Sophia Children’s Hospital Rotterdam, The Netherlands

**Keywords:** Telemedicine, frugal innovation, health system, low-resource setting, adoption, diffusion

## Abstract

**Introduction:** Access to healthcare remains a critical challenge, particularly in underserved health systems. Telemedicine, a technology-driven solution offering remote medical services, has the potential to enhance healthcare access. However, it remains primarily studied in high-resource settings and continues to face challenges in adoption and scale-up (diffusion). Insights from frugal innovation could offer valuable contributions to overcome these obstacles, promoting greater affordability and accessibility. Frugal innovation is a resource-scarce solution, typically emerging from extreme constraints, emphasizing affordability, accessibility, simplicity, efficiency, and resourcefulness. Rooted in a culturally contextualized and creative approach to problem-solving, it is particularly relevant in low-resource health systems where complex, inter-sectoral challenges occur. Despite its potential, insights from frugal innovation have not been compared, contrasted, or studied alongside telemedicine. Therefore, we will review the existing literature to analyze how both concepts and their relationships are discussed, comparing and contrasting findings. We will conduct a systematic review of empirical studies of technologies, encompassing both concepts, namely telemedicine features and frugal innovation characteristics, and explore how these features and characteristics affect adoption and diffusion pathways for the studied technologies. While we anticipate complementary aspects to arise between both concepts, our focus is to examine their reciprocal relationship and interdependence, while contributing to a more comprehensive understanding of how they impact adoption and diffusion of technology within health systems and communities in extreme resource-limited settings. Insights from our review will contribute to a deeper understanding of how to address the challenges related to the adoption and diffusion of innovative health applications in extreme resource-limited settings. Furthermore, we will advance understanding of how frugal innovation characteristics can align with traditional, non-frugal, and more advanced technologies, such as telemedicine, thereby enabling the development of innovation systems that are resource-efficient, contextually adaptive, and yet scalable.

**Methods and analysis:** Following the Preferred Reporting Items for Systematic Reviews and Meta-Analyses (PRISMA) guidelines, we will perform a systematic search across various databases, including Medline ALL, Embase, Web of Science Core Collection, Cochrane Central Register of Controlled Trials, CINAHL, PsycINFO, Scopus, Dimensions, IEEE Xplore Digital Library, EconLit, International HTA database, Latin America, the Caribbean Literature on Health Sciences (LILACS), WHO Global Index Medicus, Global Health Observatory WHO. We will consider peer-reviewed studies without language or geographic restrictions and manage extracted data using Covidence. To ensure the accuracy and quality of translations, we will implement a standardized translation protocol. The search will be conducted by an information specialist. Data synthesis will be documented via a data extraction table. Two independent reviewers will conduct screening and data extraction, checking for conflicts and consistency by a third reviewer. We will provide a narrative synthesis and ensure accessibility for non-expert audiences by avoiding jargon, explaining terms as needed, and adhering to open-access standards.

**Ethics and dissemination:** We will not collect primary data, making ethical approval unnecessary. The review findings will be shared through peer-reviewed journal publications, conferences, and stakeholder meetings.

**PROSPERO registration number:** CRD42025649418

**STRENGTHS AND LIMITATIONS OF THIS STUDY:**

- We will initiate an exploration about the interdependence between telemedicine and frugal innovation, in the context of adoption and diffusion in low-resource settings.
- We will assess features of telemedicine, e.g. remote monitoring, Electronic Medical Records (EMR) integration, prescription management, and characteristics of frugal innovation, e.g. resource efficiency, local adaptation, scalability, et cetera.
- We will include findings published without language and geographical region restrictions.
- We will include studies employing a diverse range of experimental designs, both qualitative and quantitative, with a focus on adoption and diffusion, which may lead to a high degree of heterogeneity.
- Depending on findings, we aim to conduct a Latent Class Analysis (LCA) to uncover subgroups based on features and characteristics, aiming to examine how identified classes influence the adoption and diffusion of innovation in low-resource settings.

## INTRODUCTION

### Need to reimagine telemedicine as a universally accessible and affordable solution within a socio-technical paradigm

According to the WHO, telemedicine is defined as “the delivery of health care services, where distance is a critical factor, by all healthcare professionals using information and communications technologies for the exchange of valid information for diagnosis, treatment and prevention of disease and injuries, research and evaluation, and the continuing education of health care workers, with the aim of advancing the health of individuals and communities.”^1^ Telemedicine is also embedded within the WHO Global strategy on digital health 2020-2025, relating to “the field of knowledge and practice associated with the development and use of digital technologies to improve health” and it should be “developed with principles of transparency, accessibility, scalability, replicability, interoperability, privacy, security and confidentiality.”^2^

Despite this promising nature of telemedicine, its widespread use is still challenged by varying obstacles.^3^ Barriers in high-income settings mostly encompass confidentiality issues^4^, privacy and legal concerns^5^, resistance to change^6^, workflow issues^7^, and interoperability challenges^8^. Here, the adoption of telemedicine is often facilitated by existing and functioning infrastructure, established health policies, and common access to technology. In general, it can be argued that telemedicine in high-resource settings mostly operates with a strong focus on technology, even though a successful adoption is dependent on achieving a balance within a socio-technical paradigm.

Barriers in low resource settings differ. Language barriers^9^, understanding medical jargon^10^, the need to share a single phone^11^, unawareness of technology, technically challenged staff, poor design^12^, high expectations of users^13^, and implementation costs^14^ remain key challenges. As a result, healthcare systems in low-resource settings are often hindered by infrastructural voids, financial restrictions, and socio-cultural resistance. While most studies on telemedicine have been conducted in developed countries^15^, it is essential to prioritize global health by shifting focus to low-resource settings. Frugal innovation, with its emphasis on affordable and accessible solutions, considering a culturally contextualized focus, offers potential perspectives for the outlined barriers.

According to Bhatti et al., frugal innovation is defined as “innovation under constraints” and as a conceptual model with a wide range of perspectives including “market affordability constraints”, “resource constraints or scarcity”, and “institutional voids or complexities”.^16^ As outlined by Hossain et al. “Frugal innovation is a resource scarce solution (i.e., product, service, process, or business model) that is designed and *implemented* despite financial, technological, material or other resource constraints, whereby the final outcome is *significantly* cheaper than competitive offerings (if available) and is *good enough* to meet the basic needs of customers who would otherwise remain un(der)served”.^17^

Although we recognize Hossain’s definition as comprehensive foundation for frugal innovation, we approach frugal innovation from a different perspective under the following conditions.

First, we view it in the absence of an “*implementation*” condition. Instead, we see it in an adoption context, referring to a cumulative process among stakeholders on accepting and rejecting the use of a novel technology.^18^

Second, the final outcome of frugal innovation does not need to be “*significantly*” cheaper. The term is ambiguous and poorly defined, lacking quantitative criteria, contextual clarity, and standardized usage across different fields. Instead, we reject the term as a qualifier for frugal innovation.

And third, “*good enough*” as a quality criteria is a normative and subjective terminology, fostering ambiguity and unclear connotations. Instead, we advocate for not using the term as a qualifier.

While frugal innovation encompasses multiple dimensions, from a product, service, process, business model or mindset point of view, it remains unclear to what extent this concept can be used to overcome the outlined challenges of telemedicine adoption and diffusion in low-resource contexts. Existing research on frugal innovation, outside of the scope of telemedicine, has revealed intricate findings which could offer insights for adoption of telemedicine. For instance, the more complex an innovation or the setting in which it is introduced becomes, the less likely it is to be successfully adopted, scaled up, spread, and sustained,^19,20,21,22^ which may explain the reasons behind adoption challenges of telemedicine. Complexity reduction, a core aspect of frugal innovation, could offer solutions in the interest of adoption and diffusion especially for advanced technology concepts like telemedicine.^23^ Yet, many telemedicine advancements assume that it is inherently complex and therefore not frugal.

Although not yet available in a clear evidence base, studies point to the need for exploring how features of both concepts can advance access to healthcare in low-resource settings. Few researchers like Li et al. demonstrate how frugal approaches can effectively enhance telemedicine and broadening healthcare access for underprivileged populations in less developed countries by using technology affordances in use scenarios like free telemedicine camps.^15^ These “camps” utilize frugal technologies such as videoconferencing systems deployed in mobile units like “Hospitals on Wheels”. In specific, frugal videoconferencing technology supports effective physician-patient communication over distances, offering an affordable and adaptable solution to affordable access to healthcare. In addition, Li et al.’s research reveals that patient satisfaction is influenced by aligning the “media richness” of telemedicine with factors such as diagnostic complexity and healthcare needs, emphasizing how effectively implemented frugal solutions can address diverse medical requirements in a cost-efficient manner. This indicates, that the reduction of technological complexity in combination with a social-oriented healthcare need, improves adoption and access in favor of telemedicine.

Therefore, we will investigate how the features of telemedicine correspond with the characteristics of frugal innovation. By developing a comprehensive understanding of the relationship between these two concepts, our study intends to provide insights into how new concepts could evolve, integrating attributes aligned with the social values of the target populations. A frugal-oriented telemedicine approach, focusing on complexity reduction, can recalibrate the adoption challenges by emphasizing affordability, accessibility, and scalability.

Within a broader system context, we aim to discover a deeper understanding of barriers of both concepts in relation to the overall system, rather than in isolation as standalone concepts. Hereby, we adopt a socio-technical perspective, highlighting the relational dynamics between users and technology, incorporating a multi-actor view of telemedicine, while examining human-centred design features. Our contribution will be a conceptualization of an accessible and affordable telemedicine solution by exploring the interaction between telemedicine and frugal innovation within relevant contextual parameters in low-resource settings. Therefore we will examine the following research question: How do features of telemedicine and characteristics of frugal innovation contribute towards adoption and diffusion of health applications in low-resource settings?

To identify how frugal innovation and telemedicine is interconnected, we will examine *systematic interdependencies*, a notion developed by Liu et al., referring to the interconnecting parts of a *complex system*. As outlined, “the *internal variables* of a *complex system* are rarely in-dependent of each other, as the *interactions* between the *system’s components* induce *systematic interdependencies* be-tween them.”^24^

Hereby, our research focuses on two types of *systematic interdependencies*, (i) between the socio-technical system, i.e. telemedicine with frugal innovation, and (ii) *systematic interdependencies* between telemedicine and frugal innovation within a complex system and its contextual parameters, i.e. healthcare system. While the first type can be viewed as a dependency, excluding the notion of adoption and diffusion, the second type examines how a system, i.e. telemedicine can be adopted and diffused given their interaction with contextual parameters of a broader ecosystem, i.e. healthcare system and its underlying dynamics.

We hereby define adoption as a multiphase decision-making process within the contextual parameters of a narrow social ecosystem^28^, while in contrast diffusion as the spread within a broader social ecosystem.^28,29^

Our research contributes to the scientific understanding of adoption and diffusion by conceptualizing the *systematic interdependencies* of socio-technical artefacts, such as telemedicine, as dynamic, interactive, and causal processes within complex systems. While we anticipate a difficulty in isolating *systematic interdependencies*, examining the system characteristics of frugal innovation and system features of telemedicine, we expect to reveal how a general connection between both concepts could occur.

### Foundational concepts and definitions

Our study will integrate both, modified and unmodified definitions from the existing literature.

Telemedicine is defined as a multi-dimensional concept in which medical activities are performed over time and physical distance with the support of Information Communication technology (ICT). Typically, it is designed for the exchange, provision, and/or receipt of medical care and health-related information.^25^ A practical example of telemedicine, as a health technology, is presented in telemedicine camps connecting underprivileged patients with physicians in remote cities and countries.**^Error! Bookmark not defined.^**

A health technology refers to any information or knowledge given about the concrete structure-function causality of an artefact or its components in the context of health.^26^ Mobile health applications or wearables would serve as examples.

For the purpose of our review, we define frugal innovation as a resource scarce solution, encompassing the dimension of a product, service, process, or business model. It typically emerges in low-resource settings where complex, multi-sectoral challenges prevail.

In terms of products and services, the value proposition of frugal innovation typically is more affordable and accessible, compared to competitive offerings. From a process perspective, frugal innovation emphasizes simplicity, efficiency, and resourcefulness.^27^ As for a business model perspective, frugal innovation fosters a cost-effective economic structure.

As outlined, *systematic interdependencies* are critical in our examination between telemedicine and frugal innovation concepts. We hereby define it as a mmutual influence between two or more concepts, characterized by reciprocal cause-and-effect interactions, and exchanges of actions. It reflects the interconnectedness displayed by systematic interdependencies as outlined by Liu et al.^24^

As for adoption, we define it as a multiphase decision-making process in which individuals and organizations progress through several cognitive steps, including knowledge of prior conditions, attitude formation, decision-making, implementation, and confirmation.^28^ Diffusion, unlike adoption, is an enhanced process that involves the communication of an innovation among members of a broader social ecosystem through various channels over time.^28,29^ This could include the scale from a telemedicine technology from one social ecosystem into another.

Finally, we approach Latent Class Analysis (LCA) as a statistical method that generalizes different types of regression models, where the regression coefficients vary across distinct classes.^30^ In the context of telemedicine solutions, classifications into subgroups like frugal, urban high-tech, and hybrid scalable solutions could be made, based on shared features such as affordability, accessibility, and scalability (frugal innovation characteristics).

### Objectives

In this systematic review, we aim to synthesize and critically evaluate the existing literature on telemedicine and frugal innovation. The primary research question (RQ), guiding our review is:

RQ1. How do features of telemedicine and characteristics of frugal innovation contribute towards adoption and diffusion of health applications in low-resource settings?

We used the SPIDER framework to develop this research question, as it suits qualitative research by emphasizing experiences and perceptions over interventions and outcomes. SPIDER stands for Sample, Phenomenon of Interest, Design, Evaluation, Research Type. ^32^ We will further investigate the systematic interdependencies through RQ2, RQ3, and RQ4.

RQ2. What is the systematic interdependency between telemedicine and frugal innovation?

RQ3. What is the systematic interdependency between telemedicine and contextual factors?

RQ4. What is the systematic interdependency between frugal innovation and contextual factors?

In addition to these individual *systemic interdependencies* in RQ2, RQ3, and RQ4, we will also investigate contextual factors in RQ5.

RQ5. How are contextual factors, contributing towards adoption and diffusion, outlined in each article?

A deeper understanding on the innovation trajectory on additional mechanisms and evidence-based insights will be analysed in RQ6 and RQ7.

RQ6. What is the journey of the innovation across the various cognitive steps of adoption and diffusion described in each article?

RQ7. Are there additional factors that influence the adoption and diffusion of innovations in low-resource settings?

Finally, in RQ8, quantifiable variables will be assessed based on availability. The process includes identifying relevant variables, determining the optimal number of latent classes, estimating class probabilities and regression coefficients, and interpreting the results to understand how telemedicine features align with frugal innovation characteristics.

RQ8. Can variables for Latent Class Analysis be identified?

## METHODS AND ANALYSIS

This protocol adheres to the guidelines of the Preferred Reporting Items for Systematic Reviews and Meta-Analyses Protocols (PRISMA-P).^31^ The inclusion and exclusion criteria will be guided by the SPIDER framework.^32^ Findings will be synthesized based on the following:

- Sample: Any stakeholder (individuals, groups, community, or institutions) of the health system in low-resource settings. This includes human population with limited access to healthcare infrastructure or health technology, e.g. doctors, medical center, health providers, et cetera.
- Phenomenon: Interaction between frugal innovation and telemedicine.
- Design: All research designs have been included.
- Evaluation: Assessment of barriers and facilitators leading to adoption and diffusion of frugal innovation and telemedicine.
- Research Type: Empirical studies, reviews, PhD thesis, grey literature, unindexed material, quality control studies, single case studies, multiple-site studies about telemedicine according to the WHO definition.

The paper was registered with the International Prospective Register of Systematic Reviews (PROSPERO) on February 18^th^ 2025, under the registration number CRD42025649418.

### Exclusion criteria

**Table 1.** Exclusion criteria.

| Prio | Exclusion Criteria | Description | Example |
| --- | --- | --- | --- |
| 1 | Non-eligible document types | • Review<br>• Policy documents and guidelines, e.g. WHO framework concepts<br>• Commentary<br>• Editorial working papers<br>• Posters<br>• Pre-prints: Documents that have not been reviewed yet by a scientific audience.<br>• Conference papers, where only abstract is available.<br>• Letters<br>• Written forms of communications among editors<br>• Entire books | n.a. |
| 1 | Unavailable full text | If no full text is available, document will be excluded from review. | n.a. |
| 2 | Subject incompatibility for telemedicine | If one or more of the definition following criteria for telemedicine are met, the study will be excluded.<br>Information support or management systems are excluded.<br>Interventions designed to disincentives usage behaviours without medical activities performed.<br>Medical intervention is designed for non-humans, e.g. dogs, cats, horses.<br>Electronic patient records are excluded, because they do not directly relate to medical activities performed over physical distance and time.<br>Population-based and applications that have no medical activity performed or cover preventative health and lifestyle changes. | Financing app which does not involve the notion of clinical care. |
| 2 | Subject incompatibility for frugal innovation | If none of the definition criteria for frugal innovation are met, the study will be excluded.<br>Studies that only address low-resource settings, or scarce resource settings without any frugal innovation characteristics towards the intervention are excluded. | If authors label a technology as frugal, which lacks, e.g. affordability. |
| 2 | Missing adoption or diffusion focus | If telemedicine and FI are not used in the context of adoption or diffusion. | n.a. |
| 2 | Geographical mismatch | If exclusively high-resource settings are addressed. | n.a. |

### Inclusion criteria

**Table 2.** Inclusion criteria.

| Prio | Inclusion Criteria | Description | Example |
| --- | --- | --- | --- |
| 1 | Document type | • Empirical studies<br>• Unindexed material<br>• Quality control studies<br>• Single case studies<br>• Multiple-site studies about telemedicine according to the WHO definition<br>• Book chapters | n.a. |
| 1 | Languages | • Inclusion of all languages. | n.a. |
| 1 | Telemedicine | • Any concept related to the notion of telemedicine. | e-health, telehealth, e-health, m-health, and u-health, et cetera. |
| 1 | Frugal innovation | • Any concept related to the notion of frugal innovation. |  |

### Information sources and search strategy

We developed the search strategy in collaboration with an information scientist, based on consultations with a senior library informatics expert. Table 4 outlines the expertise and roles of all authors.

**Table 3.** Expertise and roles of authors.

| Author | Role | Expertise |
| --- | --- | --- |
| CA | Lead author and guarantor of review. | PhD researcher focused on telemedicine and frugal innovation, particularly in low-resource settings. |
| PP | Co-author, screening of titles and abstracts, full text review. | PhD researcher in social sciences, focused on qualitative research and participatory methods. |
| CF | Contributing author. | Digital health expert, focused on health interventions for underserved populations. |
| CN | Co-author, Search string compilation and database extraction. | Information specialist, specialized in composing search strings used in various medical databases. |
| NKM | Contributing author. | Digital health implementation and global health expert. |
| CB | Co-author, full-text review and discussions on findings. | Frugal innovation expert, with a focus on inclusive business models and innovation management. |
| SHK | Co-author, part screening, full-text review and discussions on findings. | Health systems expert, focused on critical equipment and resources. |

To sufficiently answer the RQs, our search strategy encompasses key concepts through a series of Boolean operators (“AND”, “OR”), related to: Telemedicine, frugal innovation, adoption, diffusion, and low-resource settings.

We will implement our search strategy across the following databases: Medline ALL, Embase, Web of Science Core Collection, Cochrane Central Register of Controlled Trials, CINAHL, PsycINFO, Scopus, Dimensions, IEEE Xplore Digital Library, EconLit, International HTA database, Latin America and the Caribbean Literature on Health Sciences (LILACS), WHO Global Index Medicus, and the Global Health Observatory WHO.

### Screening and data collection process

All decisions regarding study inclusion in the review will be strictly based on the eligibility (inclusion and exclusion) criteria outlined in the Eligibility Criteria section above.

We will manage all screening records using Covidence, a web-based platform that will support the screening process (including conflict resolution), full-text review, risk of bias assessment, and data extraction.

The study selection process will involve several stages: (i) initial screening of titles and abstracts, (ii) full-text review for eligibility, and (iii) final inclusion decision for the systematic review. At each stage, we will independently and in duplicate screen records to identify potentially relevant studies and assess eligibility. Disagreements between reviewers will be resolved through discussion and consensus, and if necessary, by consulting a third reviewer from the author list, with the final decision documented in Covidence.

In line with the study selection process, data will be extracted from all records eligible for final inclusion by two independent reviewers. Reviewer disagreements will be resolved through discussion, with consensus documented in the final data extraction table. Initially, two authors will screen 5% of articles for consistency, with a third reviewer resolving conflicts. Once 10% consensus is reached, the main contributor will complete the remaining screening independently. Full-text screening will be conducted by reviewers, with bi-weekly meetings and consensus meetings for conflicts.

### Data items and review outcomes

Given the outlines research questions, we will aim to extract the following data items.

**Table 4.** Data items to extract for data extraction table.

| Relevant RQs | Data items |
| --- | --- |
| RQ1. How do features of telemedicine and characteristics of frugal innovation contribute towards adoption and diffusion of health applications in low-resource settings? | • Features of telemedicine, e.g. accessibility, technology usage.<br>• Characteristics of frugal innovation, e.g. cost effectiveness, simplicity).<br>• Use-cases and functions of telemedicine and frugal innovation.<br>• Implementation information, e.g. implementation barriers and facilitators.<br>• Adoption information, e.g. adoption barriers and facilitators.<br>• Diffusion information, e.g. diffusion barriers and facilitators. |
| RQ2. What is the systematic interdependency between telemedicine and frugal innovation? | • Relationships variables between telemedicine and frugal innovation, e.g. cost effectiveness and resource-efficient design. |
| RQ3. What is the systematic interdependency between telemedicine and contextual factors? | • Features of telemedicine, e.g. remote monitoring, examination of health conditions. |
| RQ4. What is the systematic interdependency between frugal innovation and contextual factors? | • Characteristics of frugal innovation, e.g. cost effectiveness, simplicity). |
| RQ5. How are contextual factors, contributing towards adoption and diffusion, outlined in each article? | • Implementation, adoption and diffusion factors related to technical, social, organizational, geographical, regulatory, economic, or cultural factors. |
| RQ6. What is the journey of the innovation across the various cognitive steps of adoption and diffusion described in each article? | • Direction of introduction of frugal innovation (top-down/ introduced from external systems or bottom-up/ introduced within the system.<br>• Cognitive steps performed for implementation, adoption, and diffusion within a given system. |
| RQ7. Are there additional factors that influence the adoption and diffusion of innovations in low-resource settings? | • Identified triggers for frugal innovation, e.g. profit versus problem driven.<br>• Used frameworks, theories, models or guidelines.<br>• System related resource awareness.<br>• Attributes of use-cases.<br>• Any other items that have been missed in previous RQs. |
| RQ8. Can variables for Latent Class Analysis or Principle Component Analysis be identified? | • Categorical (nominal, ordinal) or continuous, e.g. demographics, health conditions. |

### Risk of bias in studies

We will assess the methodological quality of the included studies using the appropriate critical appraisal tools provided by the Mixed-Method-Appraisal-Tool (MMAT).^33^ This tool is specifically designed to assess the methodological quality of empirical studies included in systematic reviews, incorporating various study designs. The selection of the MMAT will be based on the study design of each included study. Quality assessments will be conducted independently by two reviewers. Any discrepancies will be resolved through consultation with a third reviewer.

### Synthesis methods and evidence assessment

The extracted data from identified articles will first be summarized and synthesized in an extraction table in MS Excel. This process involves organizing the information in a structured and systematic way, improving the summary of the data. The synthesized data from the extraction table will then form the basis of the final results section of the review. A narrative summary of the findings will be prepared, integrating the outcomes from all included studies. This narrative will be aligned with the RQs to ensure a clear connection between the synthesis and the overarching goals of the review. Additionally, the narrative synthesis will be supported by tables that present the extracted data from each study in a concise and accessible format. If the available data across studies is sufficiently comparable (see RQ8), a Latent Class Analysis (LCA) will be conducted. This analysis will identify underlying patterns or subgroups within the data, providing deeper insights into the findings.

Underlying patterns could reveal how specific characteristics of frugal innovation, e.g. affordability, accessibility are interconnected with contextual factors, e.g. social norms, and thereby lead to the adoption of a telemedicine application. To ensure robustness for LCA, a sensitivity analyses will be performed. This evaluation will be carried out independently by two reviewers. In cases where disagreements arise, a third reviewer will be consulted to ensure a fair and thorough assessment.

## DISCUSSION

In our review, we aim to explore the *systematic interdependencies* between telemedicine and frugal innovation in the context of health technology adoption and diffusion within low-resource healthcare systems. Our primary aim is to analyse how both concepts potentially complement each other to overcome adoption and diffusion barriers of telemedicine in underserved communities. By bringing frugal innovation into the telemedicine conversation, we envision a pathway to developing contextually relevant applications that not only address local healthcare challenges but also empower low-resource settings with adaptable, low-cost solutions.

While our review highlights the potential synergies between these two concepts, it is also clear that a deeper understanding of their *systemic interdependencies* is required to guide future research. Both concepts are deeply influenced by cognitive processes, socio-cultural dynamics, and contextual factors. We advocate that future research will examine these dynamics from an empirical point of view to better understand how the integration of telemedicine and frugal innovation can overcome the outlines barriers.

### Limitations

This systematic literature review, the first to examine frugal innovation and telemedicine in low-resource settings, will encompass both qualitative and quantitative studies to ensure a comprehensive evidence base. Given the interdisciplinary nature of the topic, we anticipate a high level of heterogeneity. To capture evidence from all relevant fields, we will include studies in all languages and incorporate snowballing. Snowball sampling will be used in both forward and backward directions to ensure a comprehensive exploration of relevant sources. We will focus on a limited number of primary sources and prioritize key studies, while addressing the challenge of varying levels of analysis across sources.

## Data Availability

All data produced in the present study are available upon reasonable request to the authors

## Author affiliations

- Department of Multi-Actor Systems, Faculty of Technology, Policy and Management, Delft University of Technology, Delft, Netherlands
- Literature Searches Support, Dordrecht, The Netherlands

## Acknowledgements

n.a.

## Collaborators

The following coauthors and collaborators are included on behalf of Delft University of Technology and Literature Searches Support, Dordrecht, The Netherlands.

## Contributors

CA: Lead author and guarantor of review.

PP: Co-author, screening of titles and abstracts, full text review.

CF: Contributing author.

CN: Co-author, Search string compilation and database extraction.

NKM: Contributing author.

CB: Co-author, full-text review and discussions on findings.

SHK: Co-author, part screening, full-text review and discussions on findings.

The artificial intelligence tool Grammarly, was employed for proofreading grammar prior to submission of the manuscript. It was not used to create content or citations, or any other tasks that could conflict with the journal’s rules on authorship. The authors are accountable for the entirety of the manuscript.

## Funding

This research is partly funded by internal Delft University of Technology funding, including the Delft Technology Fellowship (awarded to SHK).

## Competing interest

The authors declare no potential conflicts of interest with respect to the research, authorship and/or publication of this article.

## Patient and public involvement

No patients and/or the public were involved in the design, conduct, reporting, or dissemination plans of this research.

## Patient consent for publication

Not applicable.

## Provenance and peer review

Not commissioned; externally peer reviewed.

## Supplemental material

This content has been supplied by the authors. It has not been vetted by BMJ Publishing Group Limited (BMJ) and may not have been peer-reviewed. Any opinions or recommendations discussed are solely those of the authors.

## Notes

### Competing Interest Statement

The authors have declared no competing interest.

## References

1 Ryu S. Telemedicine: opportunities and developments in Member States: report on the second global survey on eHealth. Healthcare informatics research 2012;18:153–155.

2 World Health Organization. Global Strategy on Digital Health 2020-2025. Geneva: World Health Organization; 2021.

3 Ftouni R, AlJardali B, Hamdanieh M, et al. Challenges of telemedicine during the COVID-19 pandemic: a systematic review. BMC medical informatics and decision making. 2022;22.

4 Molfenter T, Boyle M, Holloway D, et al. Trends in telemedicine use in addiction treatment. Addict Sci Clin Pract. 2015;10:1–9.

5 Petersen C, DeMuro P. al and regulatory considerations associated with use of patient-generated health data from social media and mobile health (mHealth) devices. Appl Clin Inform. 2014;6:16–26.

6 Adler G, Pritchett LR, Kauth MR, et al. A pilot project to improve access to telepsychotherapy at rural clinics. Telemed J E Health. 2014;20:83–85.

7 Plaete J, Verloigne M, Crombez G, et al. What do general practitioners think about an online tailored self-regulation programme for primary prevention. European Health Psychologist. 2014;16:889.

8 May CR, Finch TL, Cornford J, et al. Integrating telecare for chronic disease management in the community: what needs to be done? BMC Health Serv Res. 2011;11:1.

9 Serrano C, Karahanna E. The compensatory interaction between user capabilities and technology capabilities in influencing task performance: An empirical assessment in telemedicine consultations. MIS Quarterly. 2016;40:597–622.

10 Coleman C. Health literacy and clear communication best practices for telemedicine. HLRP: Health Literacy Research and Practice. 2020;4:e224–e229.

11 Bigna JJ, Noubiap JJ, Plottel CS, et al. Barriers to the implementation of mobile phone reminders in pediatric HIV care: a pre-trial analysis of the Cameroonian MORE CARE study. BMC Health Serv Res. 2014;14:1.

12 El-Mahalli AA, El-Khafif SH and Al-Qahtani MF. Successes and challenges in the implementation and application of telemedicine in the eastern province of Saudi Arabia. Perspect Health Inf Manag. 2012;9.

13 Scholl J, Syed-Abdul S, Ahmed LA. A case study of an EMR system at a large hospital in India: Challenges and strategies for successful adoption. J Biomed Inform. 2011;44:958–967.

14 Ross J, Stevenson F, Lau R, et al. Exploring the challenges of implementing e-health: a protocol for an update of a systematic review of reviews. BMJ Open. 2015;5:e006773.

15 Li X, Rai A, Krishnan G. Designing cost-effective telemedicine camps for underprivileged individuals in less developed countries: A decomposed affordance-effectivity framework. Journal of the Association for Information Systems. 2020;21:2.

16 Bhatti Y, Basu RR, Barron D, et al. Frugal innovation: Models, means, methods. Cambridge University Press. 2018.

17 Hossain M, Simula H, Halme M. Can frugal go global? Diffusion patterns of frugal innovations. Technology in Society. 2016.

18 Scott A, Pasichnyk D, Dagmara Chjojecki, et al. Optimizing adoption and diffusion of medical devices at the system level. Institute of Health Economics. 2015.

19 Cresswell K, Sheikh A. Organizational issues in the implementation and adoption of health information technology. International journal of medical informatics. 2013;82:e73–e86.

20 Atun R, de Jongh T, Secci F, et al. Integration of targeted health interventions into health systems: a conceptual framework for analysis. Health Policy Plan. 2010;25:104–111.

21 Kessler R, Glasgow RE. A proposal to speed translation of healthcare research into practice: dramatic change is needed. American journal of preventive medicine. 2011;40:637–644.

22 Margraf P. Diffusion of frugal innovation and innovativeness in low-income contexts (Doctoral dissertation). 2019.

23 Rosca E, Arnold M, Bendul JC. Business models for sustainable innovation–an empirical analysis of frugal products and services. Journal of Cleaner Production. 2017;162: S133–S145.

24 Liu Y, Slotine Y, Barabási AL. Observability of complex systems. Proceedings of the National Academy of Sciences. 2013;110:2460–2465.

25 Bashshur R, Shannon G, Krupinski E, et al. The taxonomy of telemedicine. Telemedicine and e-Health. 2011;17:484–494.

26 Lim C, Fujimoto T. Frugal innovation and design changes expanding the cost-performance frontier: A Schumpeterian approach. Research Policy. 2019;48:1016–1029.

27 Nico L, Gede SW, Manuati D. I. G. A, et al. Frugal Innovation and Bricolage: a systematic literature review. Russian Journal of Agricultural and Socio-Economic Sciences. 2024;9.

28 Rogers E, Singhal A, Quinlan M. Diffusion of Innovations. In: An Integrated Approach to Communication Theory and Research. Routledge. 2008; pp. 163 –238.

29 Nandakumar AK, Beswick J, Thomas CP, et al. Pathways Of Health Technology Diffusion: The United States And Low-Income Countries. Health Affairs. 2009;28:986–995.

30 Magidson J, Vermunt JK, Madura JP. Latent class analysis. Thousand Oaks. 2004;2:549–553.

31 Page MJ, McKenzie JE, Bossuyt PM, et al. The PRISMA 2020 statement: an updated guideline for reporting systematic reviews. BMJ. 2021;372:n71.

32 Cooke A, Smith D, Booth A. The SPIDER tool for qualitative evidence synthesis. Quality Health Research. 2012;22:1435–1443.

33 Hong QN, Fàbregues S, Bartlett G, et al. The Mixed Methods Appraisal Tool (MMAT) version 2018 for information professionals and researchers. Education for information. 2018;34: 285–291.

